# Effects of chronic angiotensin inhibition on exercise cardiovascular adaptations

**DOI:** 10.64898/2026.02.03.26345524

**Authors:** Irene Labrador-Sanchez, Alfonso Moreno-Cabañas, Lucia Gonzalez-Garcia, Diego Mora-Gonzalez, Ricardo Mora-Rodriguez, Felix Morales-Palomo

## Abstract

**Background:** Angiotensin-converting enzyme inhibitors (ACEi) and angiotensin receptor blockers (ARBs) are commonly prescribed alongside exercise to manage hypertension in individuals with metabolic syndrome (MetS). However, their potential to interfere with exercise-induced physiological adaptations remains unclear.

**Methods:** In this prospective, parallel-group study, 62 sedentary obese adults with MetS completed a 16-week supervised high-intensity interval training (HIIT) program. Participants were either chronically medicated with ACEi or ARBs (antihypertensive medication group, AHM, n=27) or a non-medicated control group (CONTROL, n=35). Primary outcomes included changes in resting and graded exercise blood pressure, MetS components, and cardiorespiratory fitness (CRF).

**Results:** Both groups exhibited significant comparable improvements (all *p time x group* > 0.05) in cardiometabolic health (MetS Z-score; AHM −0.22±0.42; CONTROL - 0.30±0.33; *p time* < 0.001) and CRF (VO_2MAX_: AHM 3.9±2.1; CONTROL 5.0±3.1 mL·kg⁻¹·min⁻¹; *p time* = 0.003). Resting blood pressure decreased similarly in both groups (Mean Arterial Pressure: AHM –4.2±8.7; CONTROL –6.5±6.3 mmHg; both *p time* = 0.005; *p time x group* > 0.05). Additionally, antihypertensive medication did not interfere with the maximal (MAP; *p* time = 0.008) and submaximal (DBP; *p* time = 0.047) blood pressure exercise responses following training with no significant *time x group* interaction (both *p* > 0.05)

**Conclusions:** Chronic treatment with angiotensin antagonist medication to treat hypertension does not restrain the effects of supervised HIIT program on improving cardiovascular function, cardiorespiratory fitness, or reducing the components of MetS. Our findings support aerobic exercise training as an effective nonpharmacological co-therapy for hypertensive patients treated with angiotensin antagonists.

## INTRODUCTION

Hypertension is one of the most prevalent chronic conditions and the leading modifiable risk factor for cardiovascular disease (CVD) ^1^. Despite the widespread availability of antihypertensive medications (AHM), the global burden of hypertension continues to increase, affecting nearly one in three adults ^2^. Current clinical guidelines ^3,4^ emphasize lifestyle interventions as a first-line strategy to prevent or delay pharmacologic therapy in individuals with elevated blood pressure (BP). However, long-term adherence to lifestyle modifications is often suboptimal, and clinicians remain cautious about delaying drug therapy in high-risk patients because of concerns about progressive target organ damage ^5^. As a result, the prescription leads to a combination of pharmacologic therapy and supervised exercise training for BP management.

Global AHM consumption has increased by approximately 43% over the past decade ^6^, with prescribing trends shifting toward angiotensin-converting enzyme inhibitors (ACEi) and angiotensin receptor blockers (ARBs), which are now preferred over diuretics as first-line agents. This shift is expected to continue following the introduction of a lower diagnostic threshold for hypertension by the American Heart Association (AHA) and the American College of Cardiology (ACC) ^7^. Indeed, these pharmacological interventions inhibit the renin-angiotensin-aldosterone system (RAAS) by preventing angiotensin II synthesis or blocking AT_1_ receptors, which play a key role ^8^. While baroceptor mechanisms govern short-term BP fluctuations, the RAAS drives both acute and chronic adjustments, playing a central role in regulating vascular tone, blood volume, and systemic resistance ^9^. Beyond BP reduction, these agents exert pleiotropic cardiovascular benefits, including improved endothelial function, reduced myocardial fibrosis, and regression of left ventricular hypertrophy, translating into enhanced outcomes across diverse cardiovascular conditions ^10^.

Regular exercise training is a cornerstone intervention for both the prevention and management of hypertension ^11,12^, with its BP-lowering effectiveness supported by large-scale meta-analysis and guidelines ^13,14^. The antihypertensive benefits of exercise are partly mediated through its interactions with key neurohumoral systems, particularly the sympathetic nervous system and the RAAS ^15^. By suppressing angiotensin II and promoting vasodilation, exercise also enhances vascular function and cardioprotection ^16^. Chronic training further reduces systemic vascular resistance ^11^ and circulating levels of angiotensin II, aldosterone, and norepinephrine ^16,17^, even in the absence of changes in plasma renin activity ^18^. Thus, exercise can directly modulate RAAS activity, thereby complementing—and potentially enhancing—the effects of antihypertensive RAAS-blocking therapies.

Growing evidence suggests that combining antihypertensive pharmacotherapy with high-intensity interval training (HIIT) may produce complementary or even synergistic effects on BP regulation. In individuals with hypertension receiving standard pharmacologic treatment, HIIT has been shown to elicit additional reductions in ambulatory BP beyond those achieved with medication alone ^18,19^. Network meta-analyses further support the efficacy of structured exercise, demonstrating that reductions in systolic blood pressure (SBP) achieved through exercise training are comparable to those observed with standard antihypertensive drug therapies ^20^. In contrast, another meta-analysis ranked angiotensin receptor blockers (ARBs) as the most effective intervention for lowering BP, with exercise appearing less potent. However, this finding was likely influenced by the overwhelming predominance of pharmacological trials (87%) included in the pooled data, which may have biased comparative estimates ^21^.

Epidemiological and clinical evidence indicate that higher levels of cardiorespiratory fitness (CRF) are associated with lower BP and reduced risk of incident hypertension ^22^. Although both pharmacologic therapy and structured exercise are known to improve cardiometabolic health, the extent to which RAAS inhibition influences exercise-induced adaptations remains unclear in individuals with cardiovascular risk factors. Evidence from healthy participants receiving ACEi suggests that pharmacologic treatment does not attenuate improvements in CRF elicited by exercise training ^23^. In addition, Chant et al.^24^ reported that AHM fails to fully control BP during exercise, suggesting that the acute vascular response to physical exertion is not adequately modulated by pharmacologic therapy. However, because that study combined multiple AHM classes, the specific effects attributable to RAAS-blocking agents cannot be clearly distinguished. To our knowledge, the impact of chronic RAAS inhibition on BP responses during exercise after structured training has not yet been fully elucidated. To address this gap, the present study examined the effect of chronic ACEi or ARBs therapy on cardiometabolic and hemodynamic adaptations to a 16-week supervised HIIT program in adults with metabolic syndrome (MetS). The working hypothesis was that RAAS inhibition would not attenuate improvements in CRF, hemodynamic function, or metabolic health.

## METHODS

### Subjects

Sixty-two middle-aged volunteers (51±12 years; 29 women and 33 men) with overweight and obesity (i.e., body mass index [BMI], 32.2±4.3 kg·m^-2^) with MetS criteria ^25^ completed the study. Participants were previously sedentary as assessed by the 7-day IPAQ (International Physical Activity Questionnaire ^26^) with less than 150 min·wk^-1^ of moderate-intensity activity ^27^. Exclusion criteria included untreated cardiovascular or renal disease, any condition associated with exercise intolerance, and the use of any pharmacological therapy targeting MetS components other than ACEi or ARBs. Upon questioning at the onset of the intervention, subjects reported a stable body weight (±1%) over the 4 months preceding the survey. All subjects provided written, witnessed, and informed consent under a protocol approved by the local Virgen de la Salud Hospital Ethics Committee of Toledo (reference #170) and in accordance with the Declaration of Helsinki. This is a substudy of a larger clinical trial assessing the effects of medication and exercise interactions in individuals with MetS (ClinicalTrials.gov Identifier: NCT03019796).

### Experimental design

This prospective, parallel-group intervention study examined the effects of exercise training in participants either treated or untreated with RAAS inhibitors (ACEi or ARBs). Participants were allocated using a non-randomized block design based on medication treatment. Recruitment, screening, intervention, and all assessments were conducted in the sequence outlined in Figure 1, following the CONSORT (Consolidated Standards of Reporting Trials) guidelines. Twenty-seven individuals with MetS receiving chronic antihypertensive therapy with ACEi or ARBs for more than six months constituted the antihypertensive medication group (AHM). Thirty-five individuals with MetS who had never received chronic pharmacologic treatment served as the control group (CONTROL; Table 1).

**Figure 1.**
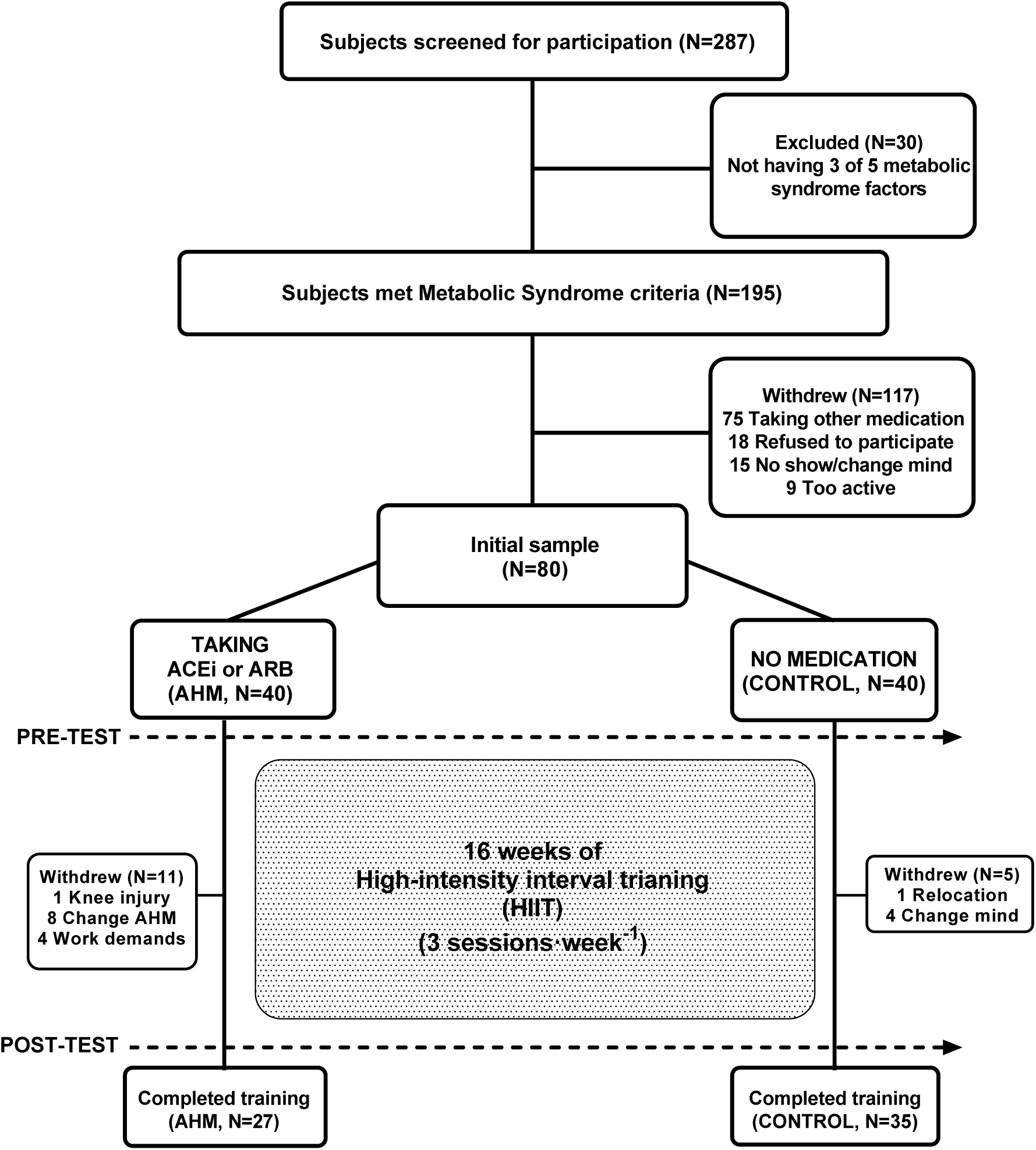
CONSORT (Consolidated Standards of Reporting Trials) schematic representation of the study procedures.

**Table 1.** Medication use.

| **Medication** | **AHM (n=27)** | **CONTROL (n=35)** |
| --- | --- | --- |
| Antihypertensive medication | 27 (100%) | - |
| **ACEi** | **12 (44%)** | - |
| Enalapril | 11 (41%) | - |
| Imidapril | 1 (4%) | - |
| **ARBs** | **15 (56%)** | - |
| Candesartan | 1 (4%) | - |
| Irbesartan | 6 (22%) | - |
| Losartan | 4 (15%) | - |
| Telmisartan | 1 (4%) | - |
| Valsartan | 3 (11%) | - |
| Cholesterol lowering | - | - |
| Triglyceride lowering | - | - |
| Glucose lowering | - | - |

Data are presented as the number of subjects taking each medication, expressed as absolute frequency (n) and proportion (%). Abbreviations: **ACEi**, angiotensin-converting enzyme inhibitors; and **ARBs**, angiotensin receptor blockers.

### Intervention

All participants completed a 16-week supervised aerobic training program, performed three times per week (Monday, Wednesday, and Friday), consisting of high-intensity interval training (HIIT) on stationary cycle ergometers (Tomahawk S-Series, Nürnberg, Germany). Participants were required to attend at least 90% of sessions to be included in the analysis. Each session began with a 10-minute warm-up at 70% of maximum heart rate (HR_MAX_), followed by four 4-minute intervals at 90% HR_MAX_, interspersed with 3-minute active recovery periods at 70% HR_MAX_, and concluded with a 5-minute cool-down. Exercise intensity was prescribed as a percentage of HR_MAX_ determined during the pre-intervention maximal exercise test. Heart rate was continuously monitored using a telemetry system (Seego, RealTrack Systems, Spain) and displayed on a large screen. Participants adjusted the cycling workload to remain within their individualized target HR zones under the direct supervision of a research team member. Training progression was implemented over the first three weeks (nine sessions), gradually increasing the number and duration of high-intensity intervals. HRMAX was reassessed monthly, and workloads were adjusted accordingly to maintain the prescribed training intensity. Participants were instructed to maintain their usual dietary and physical activity habits throughout the intervention. Monthly, they completed a 3-day dietary record (CESNID v1.0, Barcelona, Spain) and wore a wrist-based activity monitor (Polar Loop Electro, Polar, Kempele, Finland) for 48 hours. Personalized feedback was provided each month to minimize fluctuations in caloric intake and non-training physical activity.

## Outcomes measures

### Clinical investigation

Before and after the intervention, participants reported to the laboratory in the morning following an overnight fast (8–12 h). Body weight (Hawk, Mettler; Toledo, USA), height (Stadiometer, SECA 217; Hamburg, Germany), and waist circumference were measured by the same investigator according to the protocol proposed by Alberti et al. (29), using a non-elastic measuring tape. BMI was calculated, and fat mass (FM) and fat-free mass (FFM) were assessed using dual-energy X-ray absorptiometry (DXA; Hologic Discovery Wi QDR Series, Bedford, USA). Resting blood pressure was measured in triplicate using a calibrated ECG-gated automated sphygmomanometer (Tango, SunTech Medical; Morrisville, USA) after 10 minutes of supine rest. Exercise testing was performed at least 48 hours apart from fasting venous blood collection, which was used to determine serum glucose, insulin, and lipid profile (triglycerides, total cholesterol, HDL-c, and LDL-c). Insulin resistance was estimated by the homeostasis model assessment (HOMA-IR). Sex-specific Z scores were calculated for each MetS criterion using the group SD, and the sum of the Z scores for each MetS component was divided by 6 to obtain the MetS risk score. The equations used were as follows:

Men’s MetS *Z* Score= [(40 – HDL-cholesterol)/SD] + [(triglycerides – 150)/SD] + [(glucose – 100)/SD] + [(waist circumference – 94)/SD] + [(systolic blood pressure – 130)/SD] + [(diastolic blood pressure – 85)/SD]

Women’s MetS *Z* Score= [(50 – HDL-cholesterol)/SD] + [(triglycerides – 150)/SD] + [(glucose – 100)/SD] + [(waist circumference – 80)/SD] + [(systolic blood pressure – 130)/SD] + [(diastolic blood pressure – 85)/SD]

### Cardiorespiratory fitness and maximal power output

Maximal oxygen uptake (VO_2MAX_), maximal cycling power (W_MAX_), and maximal heart rate (HR_MAX_) were determined during a graded exercise test (GXT) performed on an electronically braked cycle ergometer (Ergoselect 200, Ergoline; Bitz, Germany). Gas exchange was continuously measured using indirect calorimetry (Quark RMR, Cosmed; Rome, Italy), and cardiac electrical activity was monitored via a standard 12-lead ECG (Quark T12, Cosmed; Rome, Italy). Following a 3-minute warm-up at 30 W for women and 50 W for men, the workload was increased by 15 W for women and 20 W for men every minute until volitional exhaustion. Immediately afterward, a verification test was conducted at 110% of the maximal workload achieved during the GXT to confirm attainment of VO_2MAX_ ^28^.

### Blood pressure monitoring during exercise

During the GXT, BP was manually measured by a trained practitioner using a calibrated sphygmomanometer (Gamma G7, Heine; Gilching, Germany), following established methodological standards ^29^. Measurements were obtained from the left arm, with an appropriately sized cuff positioned at heart level and the arm supported throughout the test to minimize motion artifacts and ensure accuracy. BP was recorded with participants seated on the cycle ergometer at the onset of the exercise protocol (15 W for women, 50 W for men at minute 3), and every 2 minutes at the end of each odd-numbered stage. Peak BP was determined immediately upon cessation of exercise at maximal exertion. Exercise was terminated either upon volitional exhaustion or if any predefined safety criteria for test termination were met, including chest pain with ischemic ECG changes, complex ectopy or high-grade atrioventricular block, symptomatic SBP drop >20 mmHg, severe exercise hypertension (SBP >240 mmHg or DBP >120 mmHg), oxygen desaturation, neurological symptoms, or at the discretion of the supervising physician.

### Statistical analysis

A per-protocol analysis was conducted, including only participants who completed the intervention. Sample size calculations were based on expected changes in CRF, using data from our previous study in which individuals with MetS who completed a 16-week exercise program similar to the present protocol ^30^. In this study, participants increased 0.24±0.27 L·min^-1^ following HIIT, translating to an effect size of d=1.11. Therefore, 23 participants per group should be sufficient to achieve a power of 0.95 at an alpha level of 0.05. Due to account for potential dropouts of exercise interventions (i.e., ∼30% exercise intervention attrition rates) or changing medications (primary care follow-up), the sample size was increased in each group (N=40). Data are reported as mean ± standard deviation, with 95% confidence intervals (CI) calculated for all outcome measures. Normality was verified using the Shapiro–Wilk test. Baseline comparisons between groups were performed using independent-samples t-tests. To assess the effects of training over time and between groups, a mixed-design (split-plot) ANCOVA was used for all variables, with baseline values entered as covariates. This approach allowed evaluation of time × group interactions while accounting for repeated measures (PRE and POST) within participants. Post hoc pairwise comparisons with Bonferroni correction were conducted only when significant interactions were observed. All statistical analyses were performed using SPSS v28 (IBM, Chicago, IL, USA), with significance set at *p* ≤ 0.05.

## RESULTS

### Baseline subjects’ characteristics

Participants were Caucasians living in southern Europe. Women comprise 48% of the AHM group and 46% of the CONTROL. Data were analyzed without stratification by sex, since all female participants were postmenopausal, not receiving hormone replacement therapy, and showed no significant differences compared to male participants in the main study outcomes (time x sex interaction; MetS Z score, *p* = 0.31; SBP, *p* = 0.25; DBP, *p* = 0.17; body weight, *p* = 0.31; and VO_2MAX_ (mL·Kg^-1^·min^-1^), *p* = 0.16). Subjects’ adherence to training sessions was 91% for AHM and 90% for CONTROL (*p* > 0.05). There were no differences in calorie intake or physical activity between groups. On average, subjects ingested 2296±93 at baseline and 2322±82 kcal·day^-1^ after the intervention (both *p* > 0.05). Macronutrient distribution remained stable across groups (47±5% carbohydrate, 33±2% fat [40% saturated fat], and 20±1% protein). Baseline physical activity averaged 6025±362 steps·day^-1^, 196±147 min·day^-1^ of standing, and 496±165 min·day^-1^ supine rest, with similar values after 16 weeks (587±224 steps·day^-1^, 189±77 min·day^-1^ standing and 499±201 min·day^-1^ supine rest; all *p* > 0.05).

### Body weight and composition

At baseline, participants had similar body weight, BMI, fat mass, and fat-free mass (all *p* > 0.05; Table 2). After 16 weeks of HIIT, no significant main effects of *time* or *time × group* interaction effects were observed for any anthropometric variable (all *p* > 0.05).

**Table 2.** Changes in anthropometric, MetS factors, and additional clinical variables after 16 weeks of training in both groups.

|  | AHM (n = 27) |  | CONTROL (n = 35) |  | P value |  |  |
| --- | --- | --- | --- | --- | --- | --- | --- |
|  | Baseline | 16 weeks | Baseline | 16 weeks | Baseline | Time | Time x group |
| Age (yr) | 55 ± 7 |  | 51 ± 9 |  | 0.028 |  |  |
| % Women | 48% |  | 46% |  |  |  |  |
| Anthropometric |  |  |  |  |  |  |  |
| Weight (kg) | 94.1 ± 18.5 | 93.1 ± 18.5 | 89.8 ± 15.4 | 88.2 ± 14.8 | 0.338 | 0.443 | 0.330 |
| BMI (kg·m<sup>-2</sup>) | 34.2 ± 4.7 | 33.8 ± 4.8 | 32.4 ± 3.9 | 31.8 ± 3.8 | 0.119 | 0.695 | 0.226 |
| Fat mass (kg) | 36.3 ± 10.8 | 35.6 ± 11.0 | 33.8 ± 11.6 | 32.6 ± 11.1 | 0.383 | 0.771 | 0.315 |
| Fat-free mass (kg) | 57.8 ± 13.8 | 57.4 ± 13.0 | 55.2 ± 14.9 | 54.8 ±14.7 | 0.476 | 0.057 | 0.747 |
| Metabolic Syndrome factors |  |  |  |  |  |  |  |
| Waist circumference (cm) | 109.9 ± 12.9 | 108.1 ± 13.1 | 106.4 ± 10.4 | 103.6 ± 9.4 | 0.259 | 0.110 | 0.148 |
| Glucose (mg·dL<sup>-1</sup>) | 102.1 ± 12.1 | 102.5 ± 10.4 | 105.5 ± 16.5 | 104.3 ± 18.0 | 0.366 | 0.002 | 0.899 |
| Triglycerides (mg·dL<sup>-1</sup>) | 127.1 ± 48.4 | 111.2 ± 41.8 | 149.8 ± 76.6 | 136.9 ± 72.4 | 0.165 | 0.027 | 0.350 |
| HDL-c (mg·dL<sup>-1</sup>) | 41.7 ± 8.6 | 43.0 ± 8.4 | 43.2 ± 10.9 | 43.7 ± 10.2 | 0.567 | 0.001 | 0.758 |
| MetS Z-score | 0.23 ± 0.55 | 0.01 ± 0.50 | 0.32 ± 0.61 | 0.02 ± 0.58 | 0.554 | <0.001 | 0.580 |
| Cardiometabolic risk factors |  |  |  |  |  |  |  |
| Total Cholesterol (mg·dL<sup>-1</sup>) | 192.0 ± 21.3 | 193.3 ± 28.3 | 214.4 ± 38.8 | 215.1 ± 36.9 | 0.005 | 0.096 | 0.687 |
| LDL-c (mg·dL<sup>-1</sup>) | 124.8 ± 21.3 | 128.1 ± 24.4 | 141.3 ± 33.4 | 144.0 ± 30.7 | 0.023 | 0.005 | 0.538 |
| Insulin (μIU·mL<sup>-1</sup>) | 14.4 ± 6.8 | 13.0 ± 7.2 | 12.0 ± 6.6 | 10.3 ± 4.8 | 0.161 | 0.009 | 0.303 |
| HOMA-IR | 3.7 ± 1.8 | 3.3 ± 2.0 | 3.2 ± 2.0 | 2.7 ± 1.6 | 0.360 | 0.061 | 0.405 |

Data are presented as mean ± SD.

### MetS components and additional physiological parameters

Changes in MetS components after 16 weeks of training are depicted in Table 2. Before intervention, MetS components were not different among groups (all *p* > 0.05). After intervention, we observed a significant *time* effect in glucose (AHM, 0.4; 95% CI −4.8 to 4.2 mg·dL^-1^ and CONTROL, −1.2; 95% CI −4.7 to 3.4 mg·dL^-1^; *p* = 0.002), triglycerides (AHM, −15.9; 95% CI −35.5 to −4.0 mg·dL^-1^ and CONTROL, −12.8; 95% CI −23.8 to 4.2 mg·dL^-1^; *p* = 0.027), and HDL-c (AHM, 1.3; 95% CI −1.1 to 3.3 mg·dL^-1^ and CONTROL, 0.5; 95% CI −1.3 to 2.6 mg·dL^-1^; *p* = 0.001). Similarly, the MetS Z-score showed a significant *time* effect (*p* < 0.001), with improvements of 96% in the AHM group (−0.22 SDs; 95% CI −0.37 to −0.11) and 93% in the CONTROL group (−0.30 SDs; 95% CI −0.41 to −0.17). However, no significant *time × group* interaction effects were observed for any MetS components and MetS Z-score (all *p* > 0.05). At baseline, total cholesterol (*p* = 0.005) and LDL-c (*p* = 0.023) were higher in the CONTROL than in the AHM group. Following the intervention, a significant *time* effect was observed for LDL-c (AHM, 3.2; 95% CI −4.8 to 7.8 mg·dL⁻¹ and CONTROL, 2.8; 95% CI −1.4 to 9.7 mg·dL⁻¹; *p* = 0.005) and insulin (AHM, −1.4; 95% CI −2.6 to 0.7 µIU·mL⁻¹ and CONTROL, −1.7; 95% CI −3.5 to −0.6 µIU·mL⁻¹; *p* = 0.009). However, no significant *time × group* interaction was detected for either variable (all *p* > 0.05).

### Resting blood pressure

Changes in resting BP over 16 weeks of training are shown in Figure 2. At baseline, systolic blood pressure (SBP, *p* = 0.65), diastolic blood pressure (DBP, *p* = 0.77), and mean arterial pressure (MAP, *p* = 0.69) did not differ between groups. After intervention, we observed a significant *time* effect in rest SBP (AHM, −3.2; 95% CI −7.7 to 0.6 mmHg and CONTROL, −8.0; 95% CI −11.5 to −4.2 mmHg; *p* = 0.005), DBP (AHM, −4.6; 95% CI −7.0 to −2.5 mmHg and CONTROL, −5.8; 95% CI −7.7 to −3.7 mmHg; *p* = 0.005), and MAP (AHM, −4.2; 95% CI −7.0 to −1.7 mmHg and CONTROL, −6.5; 95% CI −8.7 to −4.2 mmHg; *p* = 0.005). However, no *significant time × group* interaction effects were observed for any component of resting blood pressure (all *p* > 0.05).

**Figure 2.**
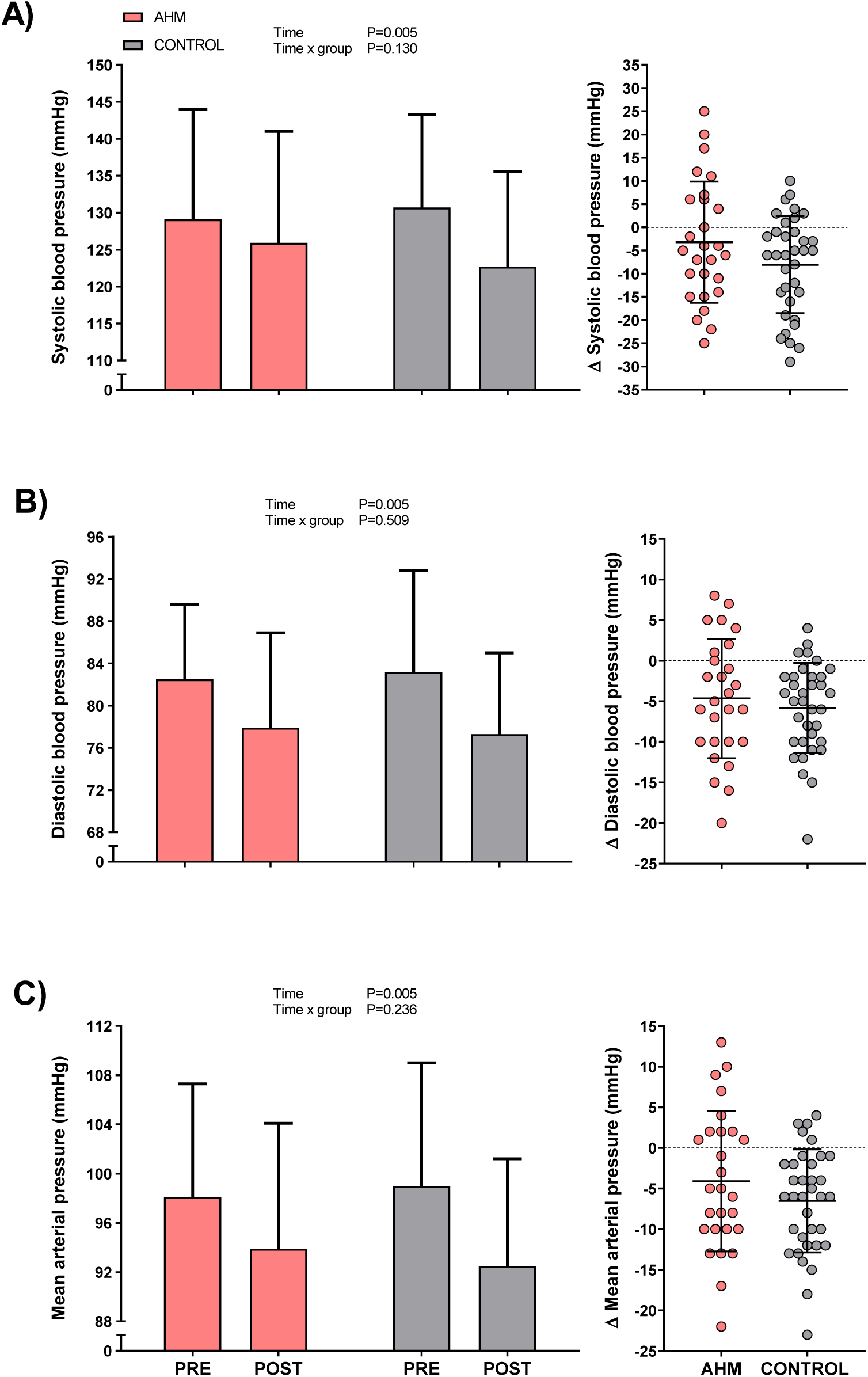
Changes in resting systolic blood pressure (A), diastolic blood pressure (B), and mean arterial pressure (C) after 16 weeks of high-intensity interval training in the AHM and CONTROL groups. Values are presented as bars and dot plots with mean ± SD.

### Cardiorespiratory fitness and maximal exercise responses

CRF (i.e., VO_2MAX_) and maximal power output (W_MAX_) evolution after intervention are shown in Figure 3. Before intervention, CRF (expressed as VO_2MAX_ in L·min^-1^ [*p* = 0.79] and mL·Kg^-1^·min^-1^ [*p* = 0.59]) and W_MAX_ (*p* = 0.41) were similar between AHM and CONTROL groups. After 16 weeks of HIIT, we observed a significant *time* effect in CRF (AHM, 0.33; 95% CI 0.24 to 0.43 L·min^-1^ and CONTROL, 0.42; 95% CI 0.34 to 0.50 L·min^-1^; *p* = 0.05) and W_MAX_ (AHM, 33; 95% CI 25 to 41 W and CONTROL, 40; 95% CI 33 to 47 W; *p* = 0.003). However, no significant *time × group* interaction effects were observed for any component of exercise performance (all *p* > 0.05). Since body weight and fat mass decreased similarly in both groups (Table 2), changes in CRF per kilogram of body weight responded similarly to the absolute values (AHM, 3.9; 95% CI 2.8 to 4.9 mL·Kg^-1^·min^-1^ and CONTROL, 5.0; 95% CI 4.1 to 5.9 mL·Kg^-1^·min^-1^; *p* = 0.003). The maximal heart rate (HR_MAX_), oxygen pulse (O_2_ Pulse at VO_2MAX_), and heart rate reserve (HRR) response are shown in Table 3. At baseline, there were no differences between groups on any of these variables (all *p* > 0.05). After exercise training, we observed a significant *time* effect in HR_MAX_ (AHM, 2; 95% CI −1 to 5 beats·min^-1^ and CONTROL, 5; 95% CI 2 to 7 beats·min^-1^; *p* < 0.001), O_2_ Pulse at VO_2MAX_ (AHM, 2.1; 95% CI 1.4 to 2.7 mL·beat^-1^ and CONTROL, 2.2; 95% CI 1.6 to 2.8 mL·beat^-1^; *p* = 0.003) and HRR (AHM, 6; 95% CI 1 to 9 beats·min^-1^ and CONTROL, 5; 95% CI 1 to 9 beats·min^-1^; *p* < 0.001).

**Figure 3.**
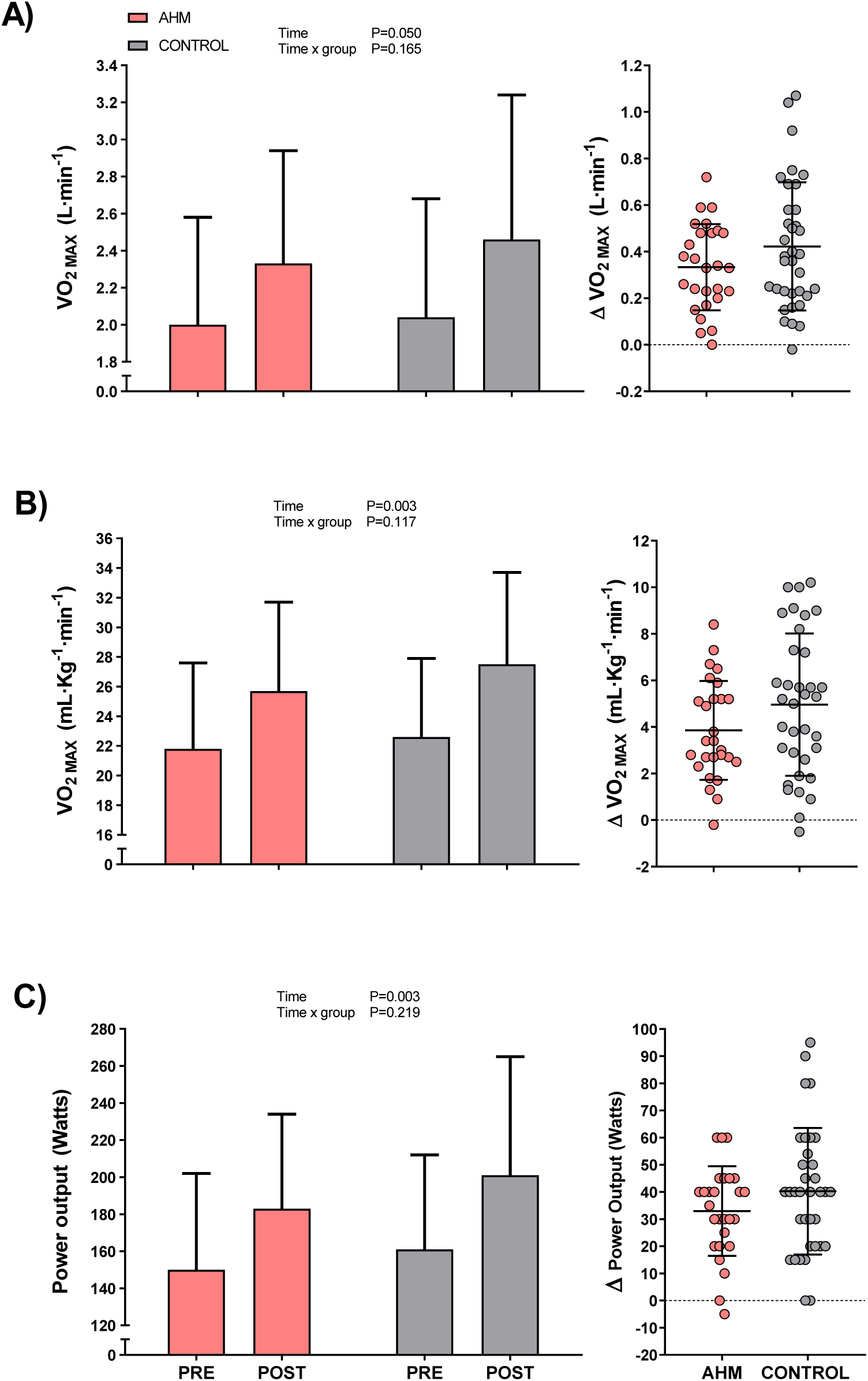
Changes in absolute VO_2MAX_ (A), relative VO_2MAX_ (B), and maximal power output (C) after 16 weeks of high-intensity interval training in the AHM and CONTROL groups. Values are presented as bars and dot plots with mean ± SD.

**Table 3.** Changes in cardiovascular and maximal exercise variables after 16 weeks of training in both groups.

|  | AHM (n = 27) |  | CONTROL (n = 35) |  | p value |  |  |
| --- | --- | --- | --- | --- | --- | --- | --- |
|  | Baseline | 16 weeks | Baseline | 16 weeks | Baseline | Time | Time x group |
| **Maximal Exercise parameters** |  |  |  |  |  |  |  |
| Maximal Heart Rate (beats·min<sup>-1</sup>) | 154 ± 15 | 156 ± 13 | 155 ± 17 | 160 ± 14 | 0.803 | **<0.001** | 0.192 |
| O<sub>2</sub> Pulse at VO<sub>2</sub>MAX (mL·beat<sup>-1</sup>) | 12.9 ± 3.5 | 15.0 ± 4.0 | 13.3 ± 5.0 | 15.5 ± 5.2 | 0.697 | **0.003** | 0.750 |
| Heart Rate Reserve (beats·min<sup>-1</sup>) | 86 ± 18 | 91 ± 17 | 88 ± 20 | 93 ± 17 | 0.635 | **<0.001** | 0.966 |
| Maximal SBP (mmHg) | 179.4 ± 22.4 | 175.5 ± 19.9 | 170.7 ± 26.4 | 171.8 ± 18.0 | 0.432 | **<0.001** | 0.961 |
| Maximal DBP (mmHg) | 93.1 ± 13.8 | 87.5 ± 12.2 | 93.6 ± 17.0 | 89.6 ± 19.1 | 0.933 | 0.138 | 0.770 |
| Maximal MAP (mmHg) | 121.8 ± 15.9 | 116.9 ± 13.5 | 119.3 ± 18.1 | 117.0 ± 15.6 | 0.743 | **0.008** | 0.806 |

Data are presented as mean ± SD. Abbreviations: SBP, Systolic Blood Pressure; DBP, Diastolic Blood Pressure; MAP, Mean Arterial Pressure.

### Blood pressure during graded exercise

BP response during the graded exercise test GXT is shown in Table 3 and Figure 4. At baseline, maximal SBP, maximal DBP, and maximal MAP did not differ between groups (all *p* > 0.05). Following the intervention, a significant main effect of *time* was observed for maximal SBP (AHM, −3.9; 95% CI −12.7 to 11.1 mmHg and CONTROL, 1.1; 95% CI −11.9 to 9.6 mmHg; *p* = 0.001) and maximal MAP (AHM, −5.0; 95% CI −13.4 to 4.9 mmHg and CONTROL, −2.3; 95% CI −11.0 to 5.4 mmHg; *p* = 0.008). However, no significant *time × group* interaction was detected for either variable (both *p* > 0.05; Table 3). Blood pressure responses at each stage of the graded exercise test—normalized to the percentage of maximal heart rate achieved (%HR_MAX_)—showed a significant main effect of *time* only for DBP (*p* = 0.047; Figure 4B) with no *time × group* interaction observed for any variable (all *p* > 0.05).

**Figure 4.**
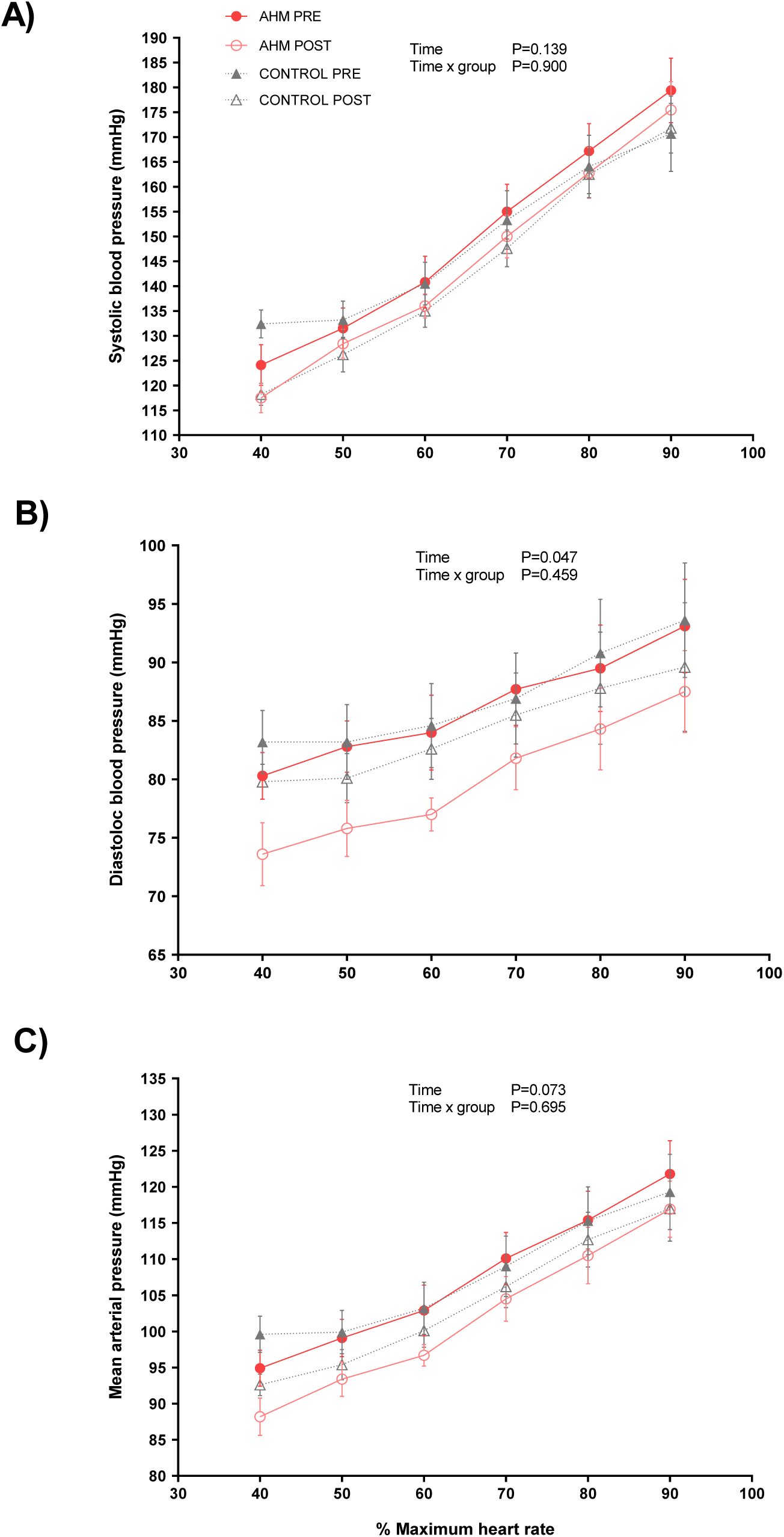
Systolic blood pressure (A), diastolic blood pressure (B), and mean arterial pressure (C) responses during graded exercise before and after 16 weeks of high-intensity interval training in the AHM and CONTROL groups. Blood pressure values are normalized to the percentage of HR_MAX_ to allow direct comparisons between groups and over time, and are presented as mean ± SD with individual data points.

## DISCUSION

In this 16-week supervised HIIT intervention, chronic treatment with ACEi or ARBs did not attenuate exercise-induced physiological adaptations in middle-aged adults with MetS. Both pharmacologically treated and untreated participants exhibited clinically meaningful improvements in blood pressure, cardiorespiratory fitness (CRF, as assessed by VO_2MAX_), and metabolic health (assessed by MetS Z-score). Importantly, no significant *time × group* interactions were detected for any cardiometabolic, hemodynamic, or performance-related outcomes, suggesting that chronic renin–angiotensin system inhibition does not impair the beneficial effects of HIIT. Additionally, body composition, dietary intake, and habitual physical activity remained stable, supporting the interpretation that observed improvements were primarily exercise-induced.

### Blood pressure reduction and cardiometabolic adaptations

The decrease in resting blood pressure observed in our participants (∼3–8 mmHg in SBP and ∼5–6 mmHg in DBP) is consistent with meta-analyses showing average decreases of ∼3–6 mmHg and ∼2–3 mmHg, respectively, after aerobic training, including HIIT ^13,14,31^. Prior evidence also indicates that structured exercise can elicit SBP reductions comparable to first-line antihypertensive therapy ^20^. Within this context, our data showed that chronic RAAS inhibition neither attenuated nor enhanced HIIT-induced adaptations, as medicated and non-medicated participants demonstrated comparable metabolic, hemodynamic, and fitness responses. These findings are consistent with placebo-controlled evidence showing that AHM and HIIT exert independent and additive effects on ambulatory BP in hypertensive individuals with MetS ^18^. Although RAAS blockade altered biochemical indices (e.g., increased renin activity and a lower aldosterone-to-renin ratio), exercise-induced BP reductions appeared to occur through mechanisms independent of RAAS suppression. In accordance with these findings, our previous work showed that only hypertensive individuals—regardless of treatment status or antihypertensive class—exhibited training-induced BP reductions, and that the magnitude of this response scaled with baseline BP, consistent with Wilder’s principle^32^.

### Acute and chronic interactions between exercise and RAAS Inhibition

Following these observations, acute studies have shown that combining a single session of aerobic exercise with AHM produces greater immediate BP reductions than either intervention alone ^19,33,34^, particularly at higher intensities ^35,36^. However, whether such acute synergistic effects persist with long-term training remains unclear. Chronic aerobic or interval training has been shown to reduce arterial stiffness, improve endothelial function ^37,38^, and decrease systemic vascular resistance ^38^ in individuals with MetS. Yet few studies have examined whether these adaptations are maintained under chronic ACEi or ARBs therapy. Our findings demonstrate that 16 weeks of HIIT elicited favorable hemodynamic and vascular adaptations in medicated participants, with no evidence that RAAS blockade interfered with these responses. Collectively, these findings support the compatibility of angiotensin antagonist medication with exercise training and reinforce the combined use of pharmacologic and lifestyle interventions in the management of hypertension and MetS.

### Cardiorespiratory fitness and clinical implications

Evidence on whether RAAS-targeting medications influence exercise-induced improvements in CRF has been controversial. Some studies in older adults have reported that ACEi therapy alone enhances exercise capacity ^39,40^ and may augment functional adaptations in frail populations ^41^. However, other trials in functionally impaired older adults ^42,43^ and healthy individuals ^23^ observed no additive effect of ACEi on training-induced gains. The present findings align with this latter evidence, demonstrating that chronic ACEi or ARBs therapy does not attenuate CRF improvements following HIIT. Absolute VO_2MAX_ increased by approximately 22% in the AHM group and 18% in the control group—equivalent to gains of ∼1.4 and ∼1.1 metabolic equivalents (METs), respectively. Exercise capacity is a strong predictor of cardiovascular and all-cause mortality in patients with hypertension ^22^. Our improvements correspond to a ∼13% reduction in all-cause mortality and a ∼15% reduction in cardiovascular events ^44^. Epidemiological data from an extensive Swedish registry indicate that individuals who increase CRF by >3% per year have an 11% lower risk of incident hypertension. In contrast, those who experience CRF declines have up to a 25% increased risk, independent of lifestyle factors ^45^. Collectively, these results suggest that HIIT-induced improvements in CRF may contribute to both short-term BP control and long-term decreases in cardiovascular risk, and that these benefits are preserved despite chronic RAAS inhibition.

### Blood pressure responses during exercise

During dynamic exercise, SBP rises proportionally with intensity due to increased cardiac output driven by sympathetic activation, whereas DBP typically remains stable or slightly decreases owing to peripheral vasodilation ^46^. Exaggerated SBP responses reflect impaired vascular regulation and are associated with arterial stiffness, endothelial dysfunction, and increased long-term cardiovascular risk ^29,47^. Chant et al. ^24^ demonstrated that individuals with treated and apparently controlled hypertension exhibit elevated SBP responses during both submaximal and maximal exercise, comparable to those observed in untreated or uncontrolled hypertension. This phenomenon has been attributed, at least in part, to heightened metaboreflex sensitivity that appears relatively resistant to conventional antihypertensive pharmacotherapy. In contrast, baseline SBP responses across exercise intensities did not differ between groups in the present study, suggesting that chronic angiotensin receptor blockade did not materially alter the acute exercise pressor response. Discrepancies with the findings of Chant et al. may be explained by differences in baseline hypertension severity and treatment burden. Notably, participants in the Chant cohort entered the study with resting SBP values near the hypertensive threshold (∼138 mmHg ^4^) and were predominantly managed with multidrug regimens, a clinical phenotype more likely to exhibit persistently exaggerated pressor responses during physical stress.

After 16 weeks of HIIT, both groups exhibited a significant *time* effect in maximal MAP and submaximal DBP (Figure 4B), accompanied by improvements in workload capacity, CRF, heart rate reserve, and maximal oxygen pulse (Figure 3, Table 3). These adaptations likely reflect enhanced left ventricular stroke volume and vascular function, consistent with evidence linking oxygen pulse kinetics to systolic and diastolic performance in older adults with mild hypertension ^48^. These results align with prior studies reporting modest reductions in exercise BP following training ^49^, and with a pooled analysis of 10 RCTs showing a ∼7 mmHg decrease in exercise SBP after aerobic training despite heterogeneity in age, BP status, and intervention protocols ^50^. Collectively, these findings suggest that HIIT enhances vascular dynamics and exercise tolerance, moderating MAP and DBP responses through improved vascular compliance, endothelial function, and attenuated sympathetic activation, even in the context of chronic RAAS inhibition.

### Strengths and Limitations

The primary strength of this study is its prospective, supervised intervention design, which enabled a direct comparison of exercise-induced adaptations between two clinically relevant populations: adults with MetS under chronic ACEi or ARBs therapy and those not receiving pharmacological treatment. This pragmatic approach enhances clinical applicability and provides insights into real-world interactions between pharmacologic RAAS inhibition and exercise-induced cardiovascular adaptations. However, several limitations should be acknowledged. First, although baseline adjustment was performed, the absence of randomization warrants caution when inferring causality. Second, pharmacologic heterogeneity—specifically the use of multiple ACEi and ARBs agents at varying doses—may have influenced vascular and hemodynamic outcomes. Future randomized controlled trials with standardized pharmacotherapy protocols are needed to delineate the specific contributions of individual RAAS inhibitors to exercise-induced adaptations.

### Conclusions

In summary, chronic ACEi or ARBs therapy did not attenuate the cardiometabolic, hemodynamic, or functional adaptations elicited by 16 weeks of supervised HIIT in adults with MetS. Medicated and non-medicated participants exhibited comparable improvements in resting and exercise blood pressure, CRF, and metabolic health, with no evidence of impaired responsiveness associated with chronic RAAS inhibition. These findings support the physiological compatibility of RAAS blockade with structured exercise training and reinforce the role of HIIT as an effective therapeutic strategy in the management of MetS among individuals receiving long-term antihypertensive treatment.

## Data Availability

All data referred to in this manuscript are available from the corresponding author upon reasonable request

## ACKNOWLEDGEMENTS

COMPETING INTERESTS AND FUNDING

Spanish Ministry of Economy, Industry and Competivity (DEP-2017-83244-R) and Spanish Ministry of Science and Innovation (PID2020-116159RB-IOO MCIN/AEI/10.13039/501100011033). The granting agencies have no role in the design, execution, or reporting of the results of this study.

## STATEMENTS AND DECLARATIONS

The authors declare that they have no competing interests.

## REFERENCES

1. Yusuf S, Joseph P, Rangarajan S, Islam S, Mente A, Hystad P, Brauer M, Kutty VR, Gupta R, Wielgosz A, et al. Modifiable risk factors, cardiovascular disease, and mortality in 155 722 individuals from 21 high-income, middle-income, and low-income countries (PURE): a prospective cohort study. The Lancet. 2020;395:795–808. doi: 10.1016/S0140-6736(19)32008-2

2. Mills KT, Stefanescu A, He J. The global epidemiology of hypertension. Nature reviews Nephrology. 2020;16:223–237. doi: 10.1038/s41581-019-0244-2

3. Arnett DK, Blumenthal RS, Albert MA, Buroker AB, Goldberger ZD, Hahn EJ, Himmelfarb CD, Khera A, Lloyd-Jones D, McEvoy JW, et al. 2019 ACC/AHA Guideline on the Primary Prevention of Cardiovascular Disease: A Report of the American College of Cardiology/American Heart Association Task Force on Clinical Practice Guidelines. Circulation. 2019;140:e596–e646. doi: doi:10.1161/CIR.0000000000000678

4. McEvoy JW, McCarthy CP, Bruno RM, Brouwers S, Canavan MD, Ceconi C, Christodorescu RM, Daskalopoulou SS, Ferro CJ, Gerdts E, et al. 2024 ESC Guidelines for the management of elevated blood pressure and hypertension: Developed by the task force on the management of elevated blood pressure and hypertension of the European Society of Cardiology (ESC) and endorsed by the European Society of Endocrinology (ESE) and the European Stroke Organisation (ESO). European Heart Journal. 2024;45:3912–4018. doi: 10.1093/eurheartj/ehae178

5. Charchar FJ, Prestes PR, Mills C, Ching SM, Neupane D, Marques FZ, Sharman JE, Vogt L, Burrell LM, Korostovtseva L, et al. Lifestyle management of hypertension: International Society of Hypertension position paper endorsed by the World Hypertension League and European Society of Hypertension. Journal of hypertension. 2024;42:23–49. doi: 10.1097/hjh.0000000000003563

6. Jayawardana S, Campbell A, Aitken M, Andersson CE, Mehra MR, Mossialos E. Global consumption patterns of combination hypertension medication: An analysis of pharmaceutical sales data from 2010-2021. PLOS global public health. 2024;4:e0003698. doi: 10.1371/journal.pgph.0003698

7. Chen R, Suchard MA, Krumholz HM, Schuemie MJ, Shea S, Duke J, Pratt N, Reich CG, Madigan D, You SC, et al. Comparative First-Line Effectiveness and Safety of ACE (Angiotensin-Converting Enzyme) Inhibitors and Angiotensin Receptor Blockers: A Multinational Cohort Study. Hypertension. 2021;78:591–603. doi: doi:10.1161/HYPERTENSIONAHA.120.16667

8. Caulfield L, Heslop P, Walesby KE, Sumukadas D, Sayer AA, Witham MD. Effect of Angiotensin System Inhibitors on Physical Performance in Older People – A Systematic Review and Meta-Analysis. Journal of the American Medical Directors Association. 2021;22:1215–1221.e1212. doi: 10.1016/j.jamda.2020.07.012

9. Forrester SJ, Booz GW, Sigmund CD, Coffman TM, Kawai T, Rizzo V, Scalia R, Eguchi S. Angiotensin II Signal Transduction: An Update on Mechanisms of Physiology and Pathophysiology. Physiological reviews. 2018;98:1627–1738. doi: 10.1152/physrev.00038.2017

10. Wei J, Galaviz KI, Kowalski AJ, Magee MJ, Haw JS, Narayan KMV, Ali MK. Comparison of Cardiovascular Events Among Users of Different Classes of Antihypertension Medications: A Systematic Review and Network Meta-analysis. JAMA Network Open. 2020;3:e1921618–e1921618. doi: 10.1001/jamanetworkopen.2019.21618

11. Cornelissen VA, Fagard RH. Effects of Endurance Training on Blood Pressure, Blood Pressure–Regulating Mechanisms, and Cardiovascular Risk Factors. Hypertension. 2005;46:667–675. doi: doi:10.1161/01.HYP.0000184225.05629.51

12. Morales-Palomo F, Ramirez-Jimenez M, Ortega JF, Lopez-Galindo PL, Fernandez-Martin J, Mora-Rodriguez R. Effects of repeated yearly exposure to exercise-training on blood pressure and metabolic syndrome evolution. Journal of hypertension. 2017;35:1992–1999. doi: 10.1097/hjh.0000000000001430

13. Cornelissen VA, Smart NA. Exercise Training for Blood Pressure: A Systematic Review and Meta-analysis. Journal of the American Heart Association. 2013;2:e004473. doi: doi:10.1161/JAHA.112.004473

14. Edwards JJ, Deenmamode AHP, Griffiths M, Arnold O, Cooper NJ, Wiles JD, O’Driscoll JM. Exercise training and resting blood pressure: a large-scale pairwise and network meta-analysis of randomised controlled trials. British journal of sports medicine. 2023;57:1317–1326. doi: 10.1136/bjsports-2022-106503

15. Kaschina E. Cross-talk between exercises and renin-angiotensin-aldosterone-system blockade in hypertension. Hypertension research: official journal of the Japanese Society of Hypertension. 2024;47:1981–1983. doi: 10.1038/s41440-024-01688-6

16. Baffour-Awuah B, Man M, Goessler KF, Cornelissen VA, Dieberg G, Smart NA, Pearson MJ. Effect of exercise training on the renin-angiotensin-aldosterone system: a meta-analysis. Journal of human hypertension. 2024;38:89–101. doi: 10.1038/s41371-023-00872-4

17. Goessler K, Polito M, Cornelissen VA. Effect of exercise training on the renin-angiotensin-aldosterone system in healthy individuals: a systematic review and meta-analysis. Hypertension research: official journal of the Japanese Society of Hypertension. 2016;39:119–126. doi: 10.1038/hr.2015.100

18. Ramirez-Jimenez M, Morales-Palomo F, Moreno-Cabañas A, Alvarez-Jimenez L, Ortega JF, Mora-Rodriguez R. Effects of antihypertensive medication and high-intensity interval training in hypertensive metabolic syndrome individuals. Scandinavian journal of medicine & science in sports. 2021;31:1411–1419. doi: 10.1111/sms.13949

19. Ramirez-Jimenez M, Morales-Palomo F, Ortega JF, Mora-Rodriguez R. Effects of intense aerobic exercise and/or antihypertensive medication in individuals with metabolic syndrome. Scandinavian journal of medicine & science in sports. 2018;28:2042–2051. doi: 10.1111/sms.13218

20. Naci H, Salcher-Konrad M, Dias S, Blum MR, Sahoo SA, Nunan D, Ioannidis JPA. How does exercise treatment compare with antihypertensive medications? A network meta-analysis of 391 randomised controlled trials assessing exercise and medication effects on systolic blood pressure. British journal of sports medicine. 2019;53:859–869. doi: 10.1136/bjsports-2018-099921

21. Noone C, Leahy J, Morrissey EC, Newell J, Newell M, Dwyer CP, Murphy J, Doyle F, Murphy AW, Molloy GJ. Comparative efficacy of exercise and anti-hypertensive pharmacological interventions in reducing blood pressure in people with hypertension: A network meta-analysis. European journal of preventive cardiology. 2020;27:247–255. doi: 10.1177/2047487319879786

22. Kokkinos P. Cardiorespiratory Fitness, Exercise, and Blood Pressure. Hypertension. 2014;64:1160-1164. doi:10.1161/HYPERTENSIONAHA.114.03616

23. Sjúrðarson T, Bejder J, Breenfeldt Andersen A, Bonne T, Kyhl K, Róin T, Patursson P, Oddmarsdóttir Gregersen N, Skoradal MB, Schliemann M, et al. Effect of angiotensin-converting enzyme inhibition on cardiovascular adaptation to exercise training. Physiological reports. 2022;10:e15382. doi: 10.14814/phy2.15382

24. Chant B, Bakali M, Hinton T, Burchell AE, Nightingale AK, Paton JFR, Hart EC. Antihypertensive Treatment Fails to Control Blood Pressure During Exercise. Hypertension. 2018;72:102–109. doi: 10.1161/hypertensionaha.118.11076

25. Alberti KG, Eckel RH, Grundy SM, Zimmet PZ, Cleeman JI, Donato KA, Fruchart JC, James WP, Loria CM, Smith SC, Jr. Harmonizing the metabolic syndrome: a joint interim statement of the International Diabetes Federation Task Force on Epidemiology and Prevention; National Heart, Lung, and Blood Institute; American Heart Association; World Heart Federation; International Atherosclerosis Society; and International Association for the Study of Obesity. Circulation. 2009;120:1640–1645. doi: 10.1161/circulationaha.109.192644

26. Craig CL, Marshall AL, Sjöström M, Bauman AE, Booth ML, Ainsworth BE, Pratt M, Ekelund U, Yngve A, Sallis JF, et al. International physical activity questionnaire: 12-country reliability and validity. Medicine and science in sports and exercise. 2003;35:1381–1395. doi: 10.1249/01.mss.0000078924.61453.fb

27. Bull FC, Al-Ansari SS, Biddle S, Borodulin K, Buman MP, Cardon G, Carty C, Chaput JP, Chastin S, Chou R, et al. World Health Organization 2020 guidelines on physical activity and sedentary behaviour. British journal of sports medicine. 2020;54:1451–1462. doi: 10.1136/bjsports-2020-102955

28. Moreno-Cabañas A, Ortega JF, Morales-Palomo F, Ramirez-Jimenez M, Alvarez-Jimenez L, Pallares JG, Mora-Rodriguez R. The use of a graded exercise test may be insufficient to quantify true changes in V̇o(2max) following exercise training in unfit individuals with metabolic syndrome. Journal of applied physiology (Bethesda, Md: 1985). 2020;129:760–767. doi: 10.1152/japplphysiol.00455.2020

29. Nayor M, Gajjar P, Murthy VL, Miller PE, Velagaleti RS, Larson MG, Vasan RS, Lewis GD, Mitchell GF, Shah RV. Blood Pressure Responses During Exercise: Physiological Correlates and Clinical Implications. Arteriosclerosis, thrombosis, and vascular biology. 2023;43:163–173. doi: 10.1161/atvbaha.122.318512

30. Morales-Palomo F, Ramirez-Jimenez M, Ortega JF, Mora-Rodriguez R. Effectiveness of Aerobic Exercise Programs for Health Promotion in Metabolic Syndrome. Medicine and science in sports and exercise. 2019;51:1876–1883. doi: 10.1249/mss.0000000000001983

31. Batacan RB, Jr., Duncan MJ, Dalbo VJ, Tucker PS, Fenning AS. Effects of high-intensity interval training on cardiometabolic health: a systematic review and meta-analysis of intervention studies. British journal of sports medicine. 2017;51:494–503. doi: 10.1136/bjsports-2015-095841

32. Mora-Rodriguez R, Ortega JF, Morales-Palomo F, Ramirez-Jimenez M, Moreno-Cabañas A, Alvarez-Jimenez L. Endurance Exercise Training reduces Blood Pressure according to the Wilder’s Principle. International journal of sports medicine. 2022;43:336–343. doi: 10.1055/a-1548-6985

33. Ramirez-Jimenez M, Fernandez-Elias V, Morales-Palomo F, Ortega JF, Mora-Rodriguez R. Intense aerobic exercise lowers blood pressure in individuals with metabolic syndrome taking antihypertensive medicine. Blood pressure monitoring. 2018;23:230–236. doi: 10.1097/mbp.0000000000000328

34. Ramirez-Jimenez M, Morales-Palomo F, Ortega JF, Mora-Rodriguez R. Post-exercise Hypotension Produced by Supramaximal Interval Exercise is Potentiated by Angiotensin Receptor Blockers. International journal of sports medicine. 2019;40:756–761. doi: 10.1055/a-0927-6957

35. Ramirez-Jimenez M, Morales-Palomo F, Pallares JG, Mora-Rodriguez R, Ortega JF. Ambulatory blood pressure response to a bout of HIIT in metabolic syndrome patients. European Journal of Applied Physiology. 2017;117:1403–1411. doi: 10.1007/s00421-017-3631-z

36. Morales-Palomo F, Ramirez-Jimenez M, Ortega JF, Pallarés JG, Mora-Rodriguez R. Acute Hypotension after High-Intensity Interval Exercise in Metabolic Syndrome Patients. International journal of sports medicine. 2017;38:560–567. doi: 10.1055/s-0043-101911

37. Mora-Rodriguez R, Ramirez-Jimenez M, Fernandez-Elias VE, Guio de Prada MV, Morales-Palomo F, Pallares JG, Nelson RK, Ortega JF. Effects of aerobic interval training on arterial stiffness and microvascular function in patients with metabolic syndrome. Journal of clinical hypertension (Greenwich, Conn). 2018;20:11–18. doi: 10.1111/jch.13130

38. Mora-Rodriguez R, Fernandez-Elias VE, Morales-Palomo F, Pallares JG, Ramirez-Jimenez M, Ortega JF. Aerobic interval training reduces vascular resistances during submaximal exercise in obese metabolic syndrome individuals. Eur J Appl Physiol. 2017;117:2065–2073. doi: 10.1007/s00421-017-3697-7

39. Hutcheon SD, Gillespie ND, Crombie IK, Struthers AD, McMurdo ME. Perindopril improves six minute walking distance in older patients with left ventricular systolic dysfunction: a randomised double blind placebo controlled trial. Heart (British Cardiac Society). 2002;88:373–377. doi: 10.1136/heart.88.4.373

40. Sumukadas D, Witham MD, Struthers AD, McMurdo ME. Effect of perindopril on physical function in elderly people with functional impairment: a randomized controlled trial. CMAJ: Canadian Medical Association journal = journal de l’Association medicale canadienne. 2007;177:867–874. doi: 10.1503/cmaj.061339

41. Buford TW, Manini TM, Hsu FC, Cesari M, Anton SD, Nayfield S, Stafford RS, Church TS, Pahor M, Carter CS. Angiotensin-converting enzyme inhibitor use by older adults is associated with greater functional responses to exercise. Journal of the American Geriatrics Society. 2012;60:1244–1252. doi: 10.1111/j.1532-5415.2012.04045.x

42. Sumukadas D, Band M, Miller S, Cvoro V, Witham M, Struthers A, McConnachie A, Lloyd SM, McMurdo M. Do ACE inhibitors improve the response to exercise training in functionally impaired older adults? A randomized controlled trial. The journals of gerontology Series A, Biological sciences and medical sciences. 2014;69:736–743. doi: 10.1093/gerona/glt142

43. Baptista LC, Machado-Rodrigues AM, Veríssimo MT, Martins RA. Exercise training improves functional status in hypertensive older adults under angiotensin converting enzymes inhibitors medication. Experimental gerontology. 2018;109:82–89. doi: 10.1016/j.exger.2017.06.013

44. Kodama S, Saito K, Tanaka S, Maki M, Yachi Y, Asumi M, Sugawara A, Totsuka K, Shimano H, Ohashi Y, et al. Cardiorespiratory fitness as a quantitative predictor of all-cause mortality and cardiovascular events in healthy men and women: a meta-analysis. Jama. 2009;301:2024–2035. doi: 10.1001/jama.2009.681

45. Holmlund T, Ekblom B, Börjesson M, Andersson G, Wallin P, Ekblom-Bak E. Association between change in cardiorespiratory fitness and incident hypertension in Swedish adults. European journal of preventive cardiology. 2021;28:1515–1522. doi: 10.1177/2047487320942997

46. Rowell LB. Human Cardiovascular Control. Oxford University Press; 1993.

47. Sabbahi A, Arena R, Kaminsky LA, Myers J, Phillips SA. Peak Blood Pressure Responses During Maximum Cardiopulmonary Exercise Testing: Reference Standards From FRIEND (Fitness Registry and the Importance of Exercise: A National Database). Hypertension. 2018;71:229–236. doi: 10.1161/hypertensionaha.117.10116

48. Lim JG, McAveney TJ, Fleg JL, Shapiro EP, Turner KL, Bacher AC, Ouyang P, Stewart KJ. Oxygen pulse during exercise is related to resting systolic and diastolic left ventricular function in older persons with mild hypertension. American heart journal. 2005;150:941–946. doi: 10.1016/j.ahj.2004.12.021

49. Barone BB, Wang NY, Bacher AC, Stewart KJ. Decreased exercise blood pressure in older adults after exercise training: contributions of increased fitness and decreased fatness. British journal of sports medicine. 2009;43:52–56. doi: 10.1136/bjsm.2008.050906

50. Pescatello LS, Franklin BA, Fagard R, Farquhar WB, Kelley GA, Ray CA. American College of Sports Medicine position stand. Exercise and hypertension. Medicine and science in sports and exercise. 2004;36:533–553. doi: 10.1249/01.mss.0000115224.88514.3a

